# Does educational attainment change the association between meat intake and obesity? A case-study from urban Brazil

**DOI:** 10.64898/2026.11.16.26368245

**Authors:** Jackson Laurentino, Poliana Palmeira, Pierre Levasseur

**Affiliations:** Nutrition and Public Health Studies and Research Group (Núcleo PENSO), Federal University of Campina Grande, Cuité, Paraíba, Brazil; Graduate Program in Science of Nutrition (PPGCN), Federal University of Paraíba, João Pessoa, Paraíba, Brazil; SADAPT, INRAE, AgroParisTech, Université Paris-Saclay, Paris, France

**Keywords:** Protein transition, obesity, meat substitution behaviors, nutritional inequality, Brazil

## Abstract

Brazil is one of the higher meat producer and consumer worldwide. Recently, the sustainability of the meat market is questioned on regard to its high contribution to climate change and public health concerns such as obesity. Hence, further public policies are likely to regulate the meat market and simultaneously reduce meat production and consumption. However, in the context of middle-income countries marked by strong inequalities such as Brazil, the nutritional effect of such market regulation is uncertain. Indeed, one can hypothesize that meat intake plays a protective role against poor nutritious food intake and weight gain among underprivileged settings. To test this hypothesis, this study analyzes household cross-sectional yearly surveys collected from 2018 to 2023 in the 27 Brazilian State capitals. As expected, multivariate estimates show education-specific associations between meat intake and obesity. Among the most educated individuals, declaring not consuming any meat the previous day before the survey is associated with lower body mass index (BMI) and obesity risk, as observed in previous epidemiological studies. By contrast, declaring consuming meat the previous day before the survey appears to be protective against weight gain among poorly educated individuals. We also observe education-specific meat substitution behaviors that help explain these findings. In facts, meat is more likely to be replaced by sweets and ready-to-eat meals among poorly educated individuals compared to more educated ones. We conclude that, everything being equals, meat market regulation policy might worsen public health concerns for under-privileged individuals. Important investments in the education sector and anti-poverty programs are thus needed to limit the raise of nutritional inequalities.

## 1. Introduction

Meat is an essential source of high-quality dietary protein and further micro-nutrients such as iron, zinc, selenium, vitamin B12, and vitamin D [1, 2]. However, the unprecedented levels of meat production and consumption worldwide highly contribute to global warming [3]. Worldwide livestock production accounts for 66% of greenhouse gas emissions emitted by the agricultural sector and is responsible for 33% of global greenhouse gas emissions [4]. Beyond environmental impacts, excessive meat consumption is also associated with obesity and non-communicable diseases including certain cancers [2, 5, 6]. In fact, as projections indicate that half of the global population will be overweight and one fourth will classified as obese by 2035, the health risks tied to such dietary habits become even more concerning [7]. Consequently, for public health and environmental concerns, there is growing interest in reducing meat intake in diets too rich in meat.

Regulating the meat market by price incentives through the implementation of taxes or quality norms appears to be a relevant public policy for reducing both meat-based public health expenditures and carbon footprint [8, 9]. Evidence shows that meat taxes consistently reduce meat consumption and related greenhouse gas emissions, especially among high meat eaters. As expected, these taxes seem particularly effective when targeting beef, which is the higher source of greenhouse gas emissions [10-12].

Nonetheless, the nutritional benefits of meat market regulations are questionable for underprivileged sub-populations. Indeed, price-based instruments like tax are known to be socially unfair, by similarly taxing the rich and the poor despite of strong differences in economic (e.g., purchasing power) and socio-cultural (e.g., health and nutrition knowledge and interest) patterns. While it is predictable that most consumers will adjust their diet in response to higher meat price, the specific changes they make (i.e., opting for healthier or unhealthier meat alternatives) are uncertain and may result in different nutritional and health outcomes depending on individual socioeconomic status (SES). Even if individuals with lower SES are usually more responsive to unhealthy food taxes due to a higher price elasticity [13, 14], substitution effects to affordable non-taxed unhealthy alternatives might also be observed [13]. One may therefore hypothesize that individuals with lower SES may be more likely to turn to energy-dense unhealthy meat alternatives (e.g., snacks, sweets and soft-drinks), leading to negative health outcomes such as weight gain and obesity.

The existing public health literature on the meat-obesity association suggests a negative association between meatless diet and obesity. However, while potential different effects according to SES are rarely explored, mots of studies are limited by sample selection biases enable to overstate the protective effects of meat less diet against obesity. While some studies simply compare nutritional outcomes of meat eaters and vegetarians [15-17], even if the latter are known to be more educated and more prone to have healthier habits than the average, others rely on non-representative cohort data mainly composed of women with a high interest for food and nutrition [18]. Consequently, one can consider that male and poorer individuals represent a black spot in the literature analyzing the meat-BMI association. To clarify this gray area in the literature, a recent study investigates potential heterogeneity in the meat-BMI association according to SES relying on a nationally representative household survey collected in France [19]. Interestingly, while they find that higher meat intake is associated with higher sugar and fat intake and higher BMI among individuals with upper SES, as demonstrated by previous studies, they reversely observe that meat intake is protective against poor nutritious food intake and weight gain among lower SES’ individuals.

SES-specific effects in the meat-BMI association might be even more concerning among developing countries characterized by substantial income inequalities. This article therefore aims to explore this potential heterogeneity in the case of urban Brazil, relying on repeated cross-sectional surveys from 2018 to 2023 collected in each State capitals. Our focus on the Brazilian adult population is relevant in several respects. Indeed, being a middle-income country with around 1/4 of its population under the poverty line, Brazil is also one of the world’s leading meat producers and consumers [20]. The place of meat is so central in the Brazilian food culture that it is consumed similarly in all social strata of the population [21]. Moreover, obesity prevalence in Brazil is worrying (22.5% in 2021), with a stronger increase among lower social groups [22]. Hence, it is expected that regulating the meat market will produce important SES-specific impacts in Brazil, with potential differences in meat substitution strategies along the social ladder. For a better control of obesity trends and the rise of nutritional inequalities, it is therefore needed to know what will be the potential nutritional effects of reducing meat in diet for all, considering potential SES-specific substitution responses.

The rest of the article is structured as follows. In Section 2, we describe the data and methods we use to estimate SES-specific associations between meat intake and BMI, controlling for several individual, household and area factors. Then, we present the results in Section 3 and finally provide a discussion in Section 4.

## 2. Methods

### 2.1 Data and sample

The study relies on the repeated cross-sectional by-phone survey Vigitel Brazil (Surveillance System for Risk and Protective Factors for Chronic Diseases by Telephone Survey) that continuously monitors the frequency and distribution of significant risk and protective factors for noncommunicable diseases (NCDs) among adults living in the 27 Brazilian State capitals quasi-annually [23, 24]. Vigitel provides probabilistic samples of the adult population (≥18 years old) residing in households with at least one landline telephone [23, 24]. Due to difficulties imposed by the COVID-19 pandemic on data collection, the Vigitel survey was canceled in 2022, and reduced (in terms of sample size) in 2020, 2021, and 2023.^1^ After merging, the repeated cross-section sample includes more than 100,000 adults who were yearly surveyed from 2018 to 2023 (except for 2022), i.e., more than 20,000 observations per survey year.

The Vigitel dataset is particularly appropriate for our research question because it has the advantage of providing across time individual information on reported food consumption (the previous day before each survey), sociodemographic characteristics (e.g., age, gender, and race/color), and anthropometric measurements (i.e., self-declared height and weight). As recommended by the survey administration, our statistical and econometric analyses are systematically balanced using the weights provided in the survey. To avoid body weight measurement errors, we restrict the sample to non-pregnant and non-lactating adult individuals and excluded adults over the age of 65 years to limit the effects of metabolic aging on weight gain. Hence, our analyses are representative of the urban adult population aged from 18 to 59 years and living in the 27 Brazilian State capitals (i.e., the 27 largest cities).

### 2.2 Estimation model

To test for specific meat substitution behaviors according to individual SES, we performed linear regressions of several nutritional outcomes on educational attainment, meat consumption, their mutual interactions, and a comprehensive set of covariates. Specifically, for each considered dependent variable, we regressed the following pooled ordinary least square (OLS) estimation model (i.e., pooling five cross-sectional surveys from 2018 to 2023):

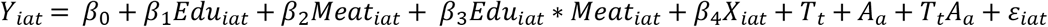

*Y*_*i*_ refers to two types of dependent outcomes that characterize the nutritional profile of an individual *i* living in a large city *a* and surveyed in the year *t*. We considered the following: (i) the individual nutritional status, measured by BMI (in kg/m^2^) and obesity status (i.e., a binary variable with a value of 1 if BMI is higher than 30 kg/m^2^, 0 otherwise); (ii) the individual consumption of food items that are commonly considered as unhealthy when excessively consumed, including sweets (e.g., industrialized flavored yogurt, sweet cakes/biscuits, desserts, ice creams, chocolates/candies, and other sugars), snacks (e.g., packaged savory snacks/chips, savory crackers or salted biscuits and other salted snacks), ready-to-eat meals (e.g., instant noodles, packaged instant soup, frozen lasagna, and other ready-to-eat frozen meals) and carbohydrates (e.g., pasta, rice, Brazilian corn couscous, and fresh corn). For each food item, the surveyed respondent must declare whether it was consumed or not the previous day before the survey.

In the set of independent variables, *Edu*_*iat*_ refers to educational attainment (in years of completed schooling) and *Meat*_*iat*_ refers to the meat eating status. We used educational attainment as proxy for individual SES for two main reasons: i) income and occupation data (i.e., other components of SES) are not available in Vigitel surveys; ii) the level of education provides a more reliable and stable image of the individual’s social position and cultural knowledge. Unlike income and occupation, educational attainment is consensually preferred in pooled cross-sectional studies to measure SES because this factor does not widely vary across time among adult populations [25].^2^ To measure meat eating status, we used a four-categories variable distinguishing respondents that declared no consuming any meat (reference category) from respondents that declared consuming only processed meat (e.g., sausages, salami, mortadella, and ham), only raw meat (e.g., beef, pork, chicken, and fish) and both raw and processed meat, the previous day before survey. To explore the association between nutritional outcomes and meat consumption according to educational attainment, we introduced an interaction term between the two factors of interest (*Edu*_*iat*_ * *Meat*_*iat*_). Using this interaction term, we could determine whether the associations between meat consumption and nutritional outcomes were significantly different across education status.

We included the main indirect determinants of nutritional outcomes in the set of control variables, *X*_*i*_, excluding direct drivers like calorie intake. Concretely, we controlled for demographic factors such as age and its square (to account for metabolic changes due to aging), sex, race/color (i.e., white, black or mixed, and other), marital status (i.e., married, single, and separated or widowed). We also controlled for individual habits including daily smoking (a binary-response variable) and physical activity practice (in minutes per day).

Finally, *T*_*t*_ and *A*_*a*_ refer to time fixed-effects (for the five survey years, ranging from 2018 to 2021 and 2023) and area fixed-effects (for the 27 State capitals of Brazil), respectively. *T*_*t*_*A*_*a*_ is their mutual interaction to neutralize baseline local specificities and potential shocks that may influence nutritional outcomes across cities and time, and make all survey waves comparable. Both fixed effects enable to deal with some forms of endogeneity, especially mitigating omitted variable bias due to time- and area-specific unobserved factors such as inflation periods, economic cycles, cultural patterns, availability of infrastructure and public services, and environmental condition.

### 2.3 Model assumptions regarding endogeneity issues

Even if our pooled time- and area-fixed effects model allows to deal with important sources of estimation bias (e.g., local inflation in meat price and other food that may simultaneously affect meat intake and nutritional outcomes), it potentially remains sensitive to other sources of endogeneity that may lead to erroneously over- or under-estimate the education-specific effects of meat intake on nutritional outcomes.

First, time- and area-fixed effects cannot mitigate individual-level omitted variable bias, given that some unobserved individual drivers of nutritional outcomes might be correlated with meat intake (e.g., food preferences and health awareness, experience in composing meatless meals, and family shocks). For instance, a positive correlation between education and healthier lifestyle and nutrition could overstate the protective effect of meatless diet on nutritional outcomes. However, it seems that health awareness are disconnected to meat intake in Brazil where meat consumption is broadly and culturally accepted and consumed in all social groups [21]. As shown in Table 1, non-meat eaters tend to be less educated and slimmer on average than meat eaters in the sample, thus rejecting the risk of an overstating bias. Another issue regarding unobserved individual heterogeneity relies on time-varying shocks or traumatizing events that may affect individuals’ diet and BMI, such as family/emotional drama (e.g., divorce, death, job loss, or income-specific inflation response). While some of these shocks may understate the nutritional differences across education levels, for instance price shocks causing weight gain through reduced purchasing power and dietary adjustments [26, 27], other shocks may overstate these differences, such as sudden illnesses that lead to both weight loss and reduced meat consumption. Unfortunately, we have no means at our disposal to control for such endogeneity sources, except for local meat inflation.^3^ Hence, we have no other choice to assume that both over- and under-stating effects compensate and neutralize each other.

**Table 1:** Sample description (means) by meat eating status, Vigitel-Brazil, 2008-2023.

|  | **No meat** | **Only processed meat** | **Only raw meat** | **Both raw and processed meat** |
|---|---|---|---|---|
|  | Mean | Mean | Mean | Mean |
| Male (dummy) | *0,439* | 0,548 | 0,45 | 0,571 |
| Age (years) | 39,002 | 37,445 | 39,704 | 37,376 |
| White race/color (dummy) | 0,406 | 0,372 | 0,402 | 0,405 |
| Black/mixed race/color (dummy) | 0,532 | 0,577 | 0,542 | 0,544 |
| Other race/color (dummy) | 0,062 | 0,052 | 0,056 | 0,051 |
| Married (dummy) | **0,517** | 0,530 | 0,451 | 0,490 |
| Single (dummy) | *0,386* | 0,398 | 0,468 | 0,438 |
| Separated/Divorced/widowed (dummy) | **0,097** | 0,071 | 0,080 | 0,072 |
| Regular smoker (dummy) | 0,095 | 0,158 | 0,081 | 0,125 |
| Physical activity (minutes per day) | *28,505* | 26,929 | 30,568 | 29,267 |
| Working (dummy) | **0,363** | 0,347 | 0,307 | 0,276 |
| Educational attainment (years) | *11,062* | 10,663 | 11,564 | 11,602 |
| Body mass index (kg/m²) | *25,970* | 26,090 | 26,543 | 26,732 |
| Obesity status (dummy) | *0,179* | 0,220 | 0,214 | 0,228 |
| Sweets intake (dummy) | **0,546** | 0,514 | 0,516 | 0,439 |
| Ready-to-eat meals intake (dummy) | 0,928 | 0,886 | 0,953 | 0,908 |
| Carbohydrates intake (dummy) | **0,237** | 0,179 | 0,122 | 0,097 |
| Snacks intake (dummy) | **0,787** | 0,688 | 0,780 | 0,716 |
| Observation | 7,029 | 1,988 | 75,754 | 24,654 |

Notes: Means are weighted using the survey recommendations to make the sample representative of large Brazilian cities. Pregnant women, children (<18 years old), and the elderly (>65 years old) were excluded. Mean-comparison tests between non meat eaters and all-type meat eaters (gathering processed and/or raw meat eaters) are shown in bold and italic when the difference is significant at the 5% level (i.e., in bold/italic when means are higher/lower among non-meat eaters). Source: Pooled Vigitel sample (2018, 2019, 2020, 2021, 2023).

Further endogeneity-related bias might be due to the presence of unobserved heterogeneity within area. For instance, individuals with higher education levels may disproportionally live in wealthier neighborhoods with abundant health services and healthier food stores (e.g., selling plant proteins), both factors being theoretically negatively correlated with meat intake and BMI [28]. Given the lack of data, we were unable to accurately disentangle the real effect of educational attainment from the effect of local amenity exposure. However, it can be reasonably assumed that, among urban adults in Brazil, educational attainment is colinear to neighborhood wealth; which renders our result relatively robust to such a source of unobserved heterogeneity. Moreover, it has been shown in the US that food supply differences across neighborhoods (in terms of products’ availability and price) contribute by only 10% to nutritional inequality between low-income and high-income households [29]. In future studies, however, it will be interesting to introduce measurements of local amenities into the model to test for potential heterogeneous effects.

Finally, potential reverse causality problems in the association between meat intake and obesity might also bias OLS regressions. A first risk of reverse causality comes from the expected gap in energy needs between larger and slimmer bodies: higher BMI implying greater calorie intake. Indeed, the basal metabolic rate (BMR) of an individual with obesity is significantly higher [30]. Hence, individuals with higher BMI may need to purchase more meat to satisfy their BMR. This point is however not a major concern in our study context insofar as there is no theoretical causal effect of educational attainment on an individual’s BMR. A more concerning risk of reverse causality though may rely on the sudden adoption of a low-meat diet by people with obesity. One can assume that individuals diagnosed by a doctor as being with obesity are more likely to receive nutritional advice recommending the reduction of fatty meat in diet. Yet, educational attainment is positively correlated with health services’ attendance. As shown by Jinnette et al. [31], individuals with higher education more frequently attend the doctor and are more willing to follow their advice. Hence, there could be some individuals with obesity that have decreased their meat purchase over the period, especially among higher education groups. Theoretically, such a reverse causality may only understate the protective nutritional effect of meat reduction in diet among higher education groups, and then understate the expected opposite nutritional effects of meat consumption according to educational attainment. Consequently, this form of reverse causality is neither a major issue in our study context.

## 3. Results

### 3.1 Descriptive statistics

Table 1 lists weighted sample means for all considered variables according to each meat eating status (i.e., no meat consumed, only consuming processed meat, only consuming raw meat, and consuming both raw and processed meat). Compared to others, individuals who declare consuming no meat (the previous day before each survey) are significantly more likely to be women, single, and professionally active, but less physically active, less educated, and have a lower body mass index (BMI) and lower risk of obesity risk. No significant differences were found in age, race/color, or smoking by meat-eating status, indicating similar distributions of these characteristics across meat-eating groups in large Brazilian cities.

In line with our hypotheses, Table 2 shows different associations between meat eating status and nutritional outcomes according to the education level (comparing lower educated individuals vs. higher educated individuals). As expected, for individuals with higher schooling, meat intake is significantly associated with higher BMI and obesity prevalence. In contrast, among less educated individuals, Table 2 does not detect significant nutritional gap across meat eating status, suggesting that meatless diet is not protective against high BMI and obesity in underprivileged settings. Table 1 provides interesting insights regarding pathways that may explain a potential lack of protective effect of meatless diet for certain sub-populations. In facts, non-meat eaters were significantly more willing to declare having consumed sweets, snacks, and carbohydrates compared to meat eaters. This latter trend may indicate risky meat substitution behaviors in the sample that are likely to affect BMI and obesity risk. We thus implement a multivariate regression model in the following subsection to determine if this trend particularly concern individuals with lower educational attainment.

**Table 2:** Mean BMI and obesity prevalence across meat eating status by education level, Vigitel-Brazil, 2008-2023.

|  | **Mean BMI (in kg/m²)** |  | **Mean obesity (%)** |  |
|---|---|---|---|---|
|  | Schooling lower than 9 years | Schooling higher than 10 years | Schooling lower than 9 years | Schooling higher than 10 years |
| No meat | 26.876 | 25.619 | 0.242 | 0.155 |
| Only processed meat | 26.366 | 25.995 | 0.241 | **0.212** |
| Only raw meat | 27.501 | **26.255** | 0.272 | **0.197** |
| Both raw and processed meat | 26.934 | **26.673** | 0.237 | **0.225** |
| Observations | 20,167 | 90,139 | 20,167 | 90,139 |

Notes: Means are weighted using the survey recommendations to make the sample representative of large Brazilian cities. Pregnant women, children (<18 years old), and the elderly (>65 years old) were excluded. Mean-comparison tests across meat eating status using non-meat eaters as reference groups are shown in bold when the difference is significant at the 5% level.
Source: Pooled Vigitel sample (2018, 2019, 2020, 2021, 2023).

### 3.2 Multivariate estimates

Table 3 reports OLS estimates of body weight outcomes (BMI and obesity risk) meat eating status, educational attainment and covariates. First, to appreciate the quality of the model specification, we present in Columns 1 and 3 regression models without interaction terms between meat status and educational attainment. These results are consistent with previous studies analyzing the sociodemographic determinants of BMI and obesity in middle-income countries such as Brazil, suggesting a general model specification of acceptable quality. In line with Streb et al. [32], who also analyzed Vigitel data, we find that being a male, a middle-age adult, single, physically inactive and working, are positively and significantly correlated with BMI and obesity risk. Similarly, using data from the two most recent waves of the Brazilian National Health Survey (2013 and 2019), Ferreira et al. [33] identified sociodemographic determinants of BMI and obesity in Brazil that align with our results.

**Table 3:** Linear regressions of body weight on meat status, educational attainment and covariates.

|  | **BMI (kg/m<sup>2</sup>)** |  | **Obesity status (dummy)** |  |
|---|---|---|---|---|
|  | (1) | (2) | (3) | (4) |
| Male (dummy) | 0.380*** | 0.378*** | 0.004 | 0.004 |
|  | (0.077) | (0.077) | (0.006) | (0.006) |
| Age (years) | 0.402*** | 0.404*** | 0.019*** | 0.019*** |
|  | (0.018) | (0.018) | (0.001) | (0.001) |
| Squared age (years) | -0.004*** | -0.004*** | -0.000*** | -0.000*** |
|  | (0.000) | (0.000) | (0.000) | (0.000) |
| *Race/color (ref. White race/color)* |  |  |  |  |
| Black/mixed race/color (dummy) | 0.379*** | 0.384*** | 0.013** | 0.013** |
|  | (0.081) | (0.081) | (0.006) | (0.006) |
| Other race/color (dummy) | 0.219 | 0.214 | 0.007 | 0.006 |
|  | (0.158) | (0.157) | (0.013) | (0.013) |
| *Marital status (ref. Married)* |  |  |  |  |
| Single (dummy) | 0.679*** | 0.674*** | 0.027*** | 0.027*** |
|  | (0.088) | (0.088) | (0.007) | (0.007) |
| Separated/Divorced/widowed (dummy) | 0.458*** | 0.448*** | 0.003 | 0.003 |
|  | (0.140) | (0.138) | (0.010) | (0.010) |
| Regular smoker (dummy) | -0.986*** | -0.979*** | -0.057*** | -0.057*** |
|  | (0.130) | (0.130) | (0.010) | (0.010) |
| Physical activity (minutes per day) | -0.010*** | -0.010*** | -0.001*** | -0.001*** |
|  | (0.001) | (0.001) | (0.000) | (0.000) |
| Working (dummy) | 0.156* | 0.154* | 0.019*** | 0.019*** |
|  | (0.087) | (0.087) | (0.006) | (0.006) |
| Educational attainment (years) | -0.040*** | -0.108*** | -0.003*** | -0.007*** |
|  | (0.009) | (0.031) | (0.001) | (0.002) |
| *Meat status (ref.= no meat intake)* |  |  |  |  |
| Only processed meat (dummy) | 0.121 | -1.517 | 0.040* | -0.067 |
|  | (0.301) | (1.123) | (0.021) | (0.072) |
| Only raw meat (dummy) | 0.486*** | -0.095 | 0.032*** | -0.003 |
|  | (0.137) | (0.420) | (0.010) | (0.032) |
| Both raw and processed meat (dummy) | 0.787*** | -0.601 | 0.052*** | -0.032 |
|  | (0.149) | (0.454) | (0.011) | (0.035) |
| ***Interaction terms*** |  |  |  |  |
| **ProcessedmeatXEducation** |  | **0.151\*** |  | **0.010\*\*\*** |
|  |  | **(0.092)** |  | **(0.006)** |
| **RawmeatXEducation** |  | **0.053\*\*\*** |  | **0.003\*\*\*** |
|  |  | **(0.033)** |  | **(0.003)** |
| **BothmeatsXEducation** |  | **0.123\*\*\*** |  | **0.007\*\*\*** |
|  |  | **(0.035)** |  | **(0.003)** |
| Time fixed effect (dummies) | YES | YES | YES | YES |
| City fixed effect (dummies) | YES | YES | YES | YES |
| Interacted fixed effects | YES | YES | YES | YES |
| Constant | 16.945*** | 17.669*** | -0.232*** | -0.187*** |
|  | (0.420) | (0.567) | (0.031) | (0.041) |
| Observations | 109,425 | 109,425 | 109,425 | 109,425 |
| R-squared | 0.078 | 0.079 | 0.038 | 0.038 |

Notes: Estimates are weighted using the survey recommendations to guarantee the representativeness of the sample. Pregnant women, children (<18 years old), and the elderly (>65 years old) were excluded. Standard errors in parentheses. Significance levels are: \*\*\* p<0.01, \*\* p<0.05, \* p<0.1. Source: Pooled Vigitel sample (2018, 2019, 2020, 2021, 2023).

In contrast, having higher level of education is negatively and significantly correlated with BMI and obesity, which illustrates the social gradient of obesity in urban Brazil. This social gradient was evidenced by Ferreira et al. [33] and Silva et al. [34] who showed that higher levels of education are negatively associated with BMI and obesity risk, particularly among women, while disparities related to skin color further highlight the socio-demographic inequalities of obesity in Brazil.

Regarding meat eating status, as observed by Apreline et al. [21], Khodayari et al.[35], and Luan et al. [36], we also observe that individuals declaring having consumed only raw meat and both raw and processed meat tend to have higher BMI and obesity risk.

Columns 2 and 4 replicate the same regression model by adding interaction terms between meat eating status and educational attainment (in years) to investigate SES-specific associations. As expected, educational status appears to moderate the association between meat consumption and bodyweight. Compared to individuals who declare consuming no meat at the previous day before survey, each additional year of education increases BMI by 0.12kg/m^2^ and obesity risk by 0.7 percentage point among raw and processed meat eaters. Figure 1 plots marginal effects from Table 3, Columns 2. The figure clearly illustrates opposite associations according to educational status. While the most educated individuals tend to be slimmer when declaring not consuming meat a day before survey (with a mean BMI around 25kg/m^2^), the opposite is true for poorly educated individuals for which not consuming meat a day before the survey is associated with higher BMI (more than 27kg/m^2^ on average). Likewise, Figure 1 also shows that consuming meat a day before survey appears to be relatively protective against high BMI among poorly educated individuals while a risk factor for more educated individuals. In alternative regressions, reported Table S2 of Supplementary Materials, we also explore potential heterogeneity according to gender, replicating the OLS model for men and women specific sub-samples. We find that opposite meat-BMI associations observed according to educational attainment are particularly true for men. For women, no significant difference was detected, suggesting that the meat-BMI association remains negative independently of schooling years.

**Figure 1:**
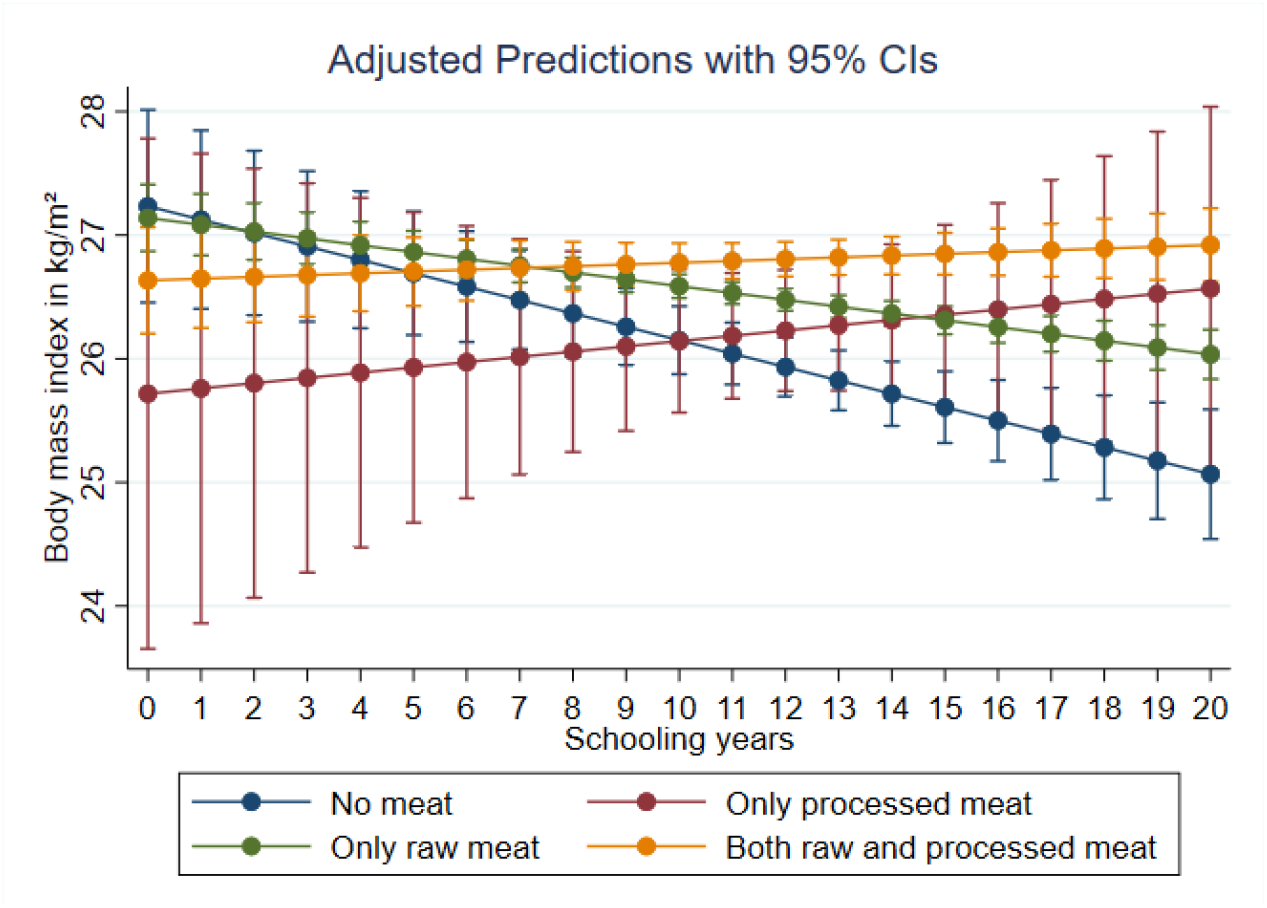
Adjusted marginal effects of educational attainment on body mass index by meat eating status. Notes: Estimates are weighted using the survey recommendations to guarantee the representativeness of the sample. Lactating and pregnant women, children (<18 years old), and the elderly (>65 years old) were excluded. Estimates are adjusted by age, gender, race/color, marital status, smoking, physical activity, and time- and area-fixed effects and their interaction. Confidence intervals (CIs) are fixed at the 95% level. Source: Pooled Vigitel sample (2018, 2019, 2020, 2021, 2023).

To analyze meat substitution pathways enable to explain a potential weight gain among poorly educated individuals in a context of meat reduction in diet, we investigate potential heterogeneous effects in the association between meat eating status and the consumption of several unhealthy food/beverage items (i.e., sweets, snacks, ready-to-eat meals, soft-drinks, carbohydrates, and ultra-processed breads) according to educational attainment. Related regressions are reported in Table S3 of Supplementary Materials. We specifically detect two main groups of unhealthy food significantly used by poorly educated individuals as meat substitutes: sweets and ready-to-eat meals. As illustrated by plotted marginal effects in Figure 2, while there is no significant difference regarding sweets and ready-to-eat meals for upper educated individuals between meat eating statuses, there are strong gaps among poorly educated individuals. Among the latter, declaring no meat consumption is associated with a larger probability to declare consuming sweets (around 70%) and ready-to-eat meals (around 90%). In contrast, when declaring consuming both types of meat, the probability to declare consuming sweets and ready-to-eat meals significantly falls (respectively around 50% and 80%).

**Figure 2:**
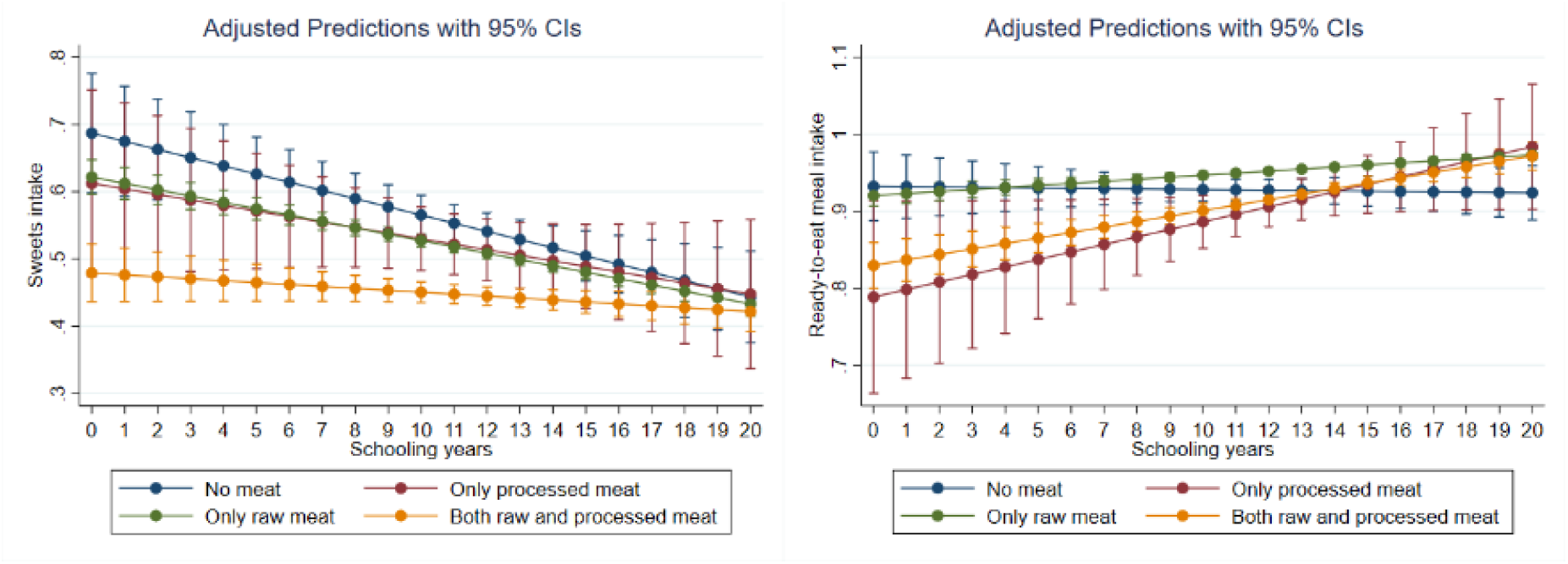
Exploring potential education-based meat substitution strategies (adjusted marginal effects) Notes: Estimates are weighted using the survey recommendations to guarantee the representativeness of the sample. Lactating and pregnant women, children (<18 years old), and the elderly (>65 years old) were excluded. Estimates are adjusted by age, gender, race/color, marital status, smoking, physical activity, city dummies, and survey years dummies. Confidence intervals (CIs) are fixed at the 95% level. Source: Pooled Vigitel sample (2018, 2019, 2020, 2021, 2023)

## 4. Discussion

In Brazil, meat consumption is determined by important factors such as purchasing power, food availability and affordability, rising prices and social inequalities [37-39]. SES is a crucial determinant of food access in Brazil [38]. Individuals in higher-income strata consume more meat, as shown by population-based studies in Brazil, where individuals with higher economic status reported higher total meat [40,41], processed meat [42,43,44], and red meat consumption [44,45]. As well, Ferreira et al. showed that food insecurity correlates with lower meat intake, noting that severe food insecurity reduced red meat consumption by 26.9% compared to food security [45]. Nevertheless, meat intake still exceeded EAT-Lancet recommendations across all income levels and food security statuses [45], a trend linked to the deep cultural integration of red meat in Brazil [46,47].

Based on a representative sample of the Brazilian adult population, this study showed educational attainment trends regarding the association between meat consumption and obesity. Although meat consumption the day before the survey was positively associated with BMI and obesity in groups with higher levels of education, our results show that meat intake is associated with lower BMI among poorly educated individuals. Consistent with expectations, this study suggests that educational status acts as a moderating variable in the link between meat consumption and body weight. To our knowledge, this study is the first to show opposite relationships between meat consumption and BMI in lower- and higher-educational-attainment groups in the Brazilian context, converging with recent results found in high-income countries like France [19]. Another important finding of the study is that meat-substitution behaviors may also depend on educational attainment. Among poorly educated individuals, meat is more likely to be replaced by sweets and ready-to-eat meals than among more educated individuals.

Our analyses also revealed another issue in the relationship between meat consumption and education level: possible health risky meat substitutes among individuals with lower education levels in the sample. Our results estimated a tendency to replace meat with ultra-processed foods (sweets and ready-to-eat meals) among individuals with lower education levels who reported not consuming meat the day before the survey. Replacing meat, a source rich in high-quality protein, iron, and zinc, with ultra-processed foods (high in sugars, saturated fat, and additives) has serious consequences. This type of substitution can accelerate weight gain and increase the risk of obesity [48]. Consuming UPFs raises the population-level risk of developing type 2 diabetes, hypertension, metabolic syndrome, and cardiovascular diseases, particularly among vulnerable segments of society [49]. This also exacerbates social and health disparities, as groups with lower SES are driven to choose UPFs due to their lower cost and availability, thereby exposing this population to the development of non-communicable chronic diseases [50,51].

In Brazil, previous studies have shown consistent increases in the prices of healthy foods and decreases in the prices of UPFs [52,53]. Caldeira at al. showed that Brazil has seen prices of ultra-processed foods fall between 2018 and 2024, and they are projected to become the cheapest food group by 2026 [54]. scenario hinders the adoption of healthy, sustainable diets, particularly among lower-SES populations [54].

Popkin et al. [55] discuss the need for robust public policies that make healthy foods more affordable and competitive than UPFs. These policies include taxing UPFs and subsidizing fruits, vegetables, and other fresh foods. Some countries now try to implement this approach [55,56]. Pereda et al. [57] report that policies combining UPF taxation with subsidies for unprocessed and minimally processed foods are fairer, as they help reduce inequalities in access to healthy foods and minimize potential negative impacts on the well-being of individuals of low SES.

Regarding environmental concerns, Cruz et al. [58] showed that both beef and UPFs had a significant linear association with carbon and water footprints in crude and adjusted models. The authors also noted that the greatest reduction in environmental footprints would occur when both beef and UPFs purchases are decreased, resulting in a 21.1% reduction in carbon footprint and a 20% reduction in water footprint. However, evidence from a recent modeling study in Brazil shows that healthy, sustainable diets are currently more expensive than the average diet, despite their benefits for health and the environment [59].

From this perspective, enhancing food sustainability in contexts of high social inequalities is a great challenge. As suggested by our results, under-privileged individuals may replace meat with affordable ultra-processed foods conducting to aggravate health inequalities. Complementary findings reported that meat costs for socially vulnerable population groups may affect consumption of other food groups, such as fruits and vegetables [40]. In this context, social inequalities complicate a consensus on meat consumption recommendations for the Brazilian population that are nutritionally, culturally, ecologically, and socially appropriate [40].

## 5. Policy implications

Developing national dietary recommendations based on sustainability is challenging in contexts of strong social inequalities, since restricting meat consumption may increase health inequalities and obesity concerns among vulnerable groups. To promote healthier, more sustainable diets, public health interventions must account for regional and socioeconomic disparities, as well as the cultural significance of meat. Policies should prioritize the human right to food and consider the socioeconomic and environmental factors that influence and limit access to meat.

The Dietary Guidelines for the Brazilian Population have sustainability as one of their guiding principles. These Guidelines recommend prioritizing natural or minimally processed foods over ultra-processed foods [59]. In this regard, public efforts should aim to increase the physical and economic availability of unprocessed and minimally processed foods, especially in vulnerable areas. Strengthening family farming and discouraging ultra-processed foods through fiscal measures are important for creating food environments that promote health and sustainability.

## 6. Limitations and strengths

The findings of this study should be analyzed in light of certain limitations. The first limitation is the cross-sectional nature of the data, which does not allow the identification of causal effects. Another limitation of this study is the self-reported nature of consumption frequency, a characteristic of the Vigitel survey. As well, the Vigitel survey does not provide income and food security data; consequently, educational attainment served as the primary proxy for SES, which may not fully capture economic hardship or food access. Despite such methodological limitations, the study highlights the importance of using large national databases, such as the Vigitel survey, to conduct comprehensive and representative analyses. In this sense, this is one of the first national-sample studies addressing educational inequalities and meat consumption in Brazil.

This study offers important insights into the relationship between education level and meat consumption. Our results have broader implications for public health in the country, including the development of public policies and targeted health interventions to reduce socioeconomic inequalities and promote access to healthy, sustainable foods.

Further studies should evaluate meat substitution strategies and the motivations behind them in greater depth. Research also should simulate and evaluate the potential impact of fiscal policies, particularly targeted taxation on ultra-processed foods, including ultra-processed meat products, to assess their effectiveness in shifting consumer behavior, improving population diet quality, and mitigating socio-economic and health disparities.

## Data Availability

The data set is publicly available at: https://www.gov.br/saude/pt-br/composicao/svsa/inqueritos-de-saude/vigitel

## Acknowledgment

The authors are grateful to the TERREE doctoral grant from the Embassy of France in Brazil that enabled to realize this study. The authors have no conflict of interest to declare.

## Footnotes

1 We chose that period because it is the years during which Vigitel added questions about processed meat consumption; previously, the Vigitel study had only questions about raw meat.

2 By contrast, household income and, to a lesser extent, individual occupation might be preferred in longitudinal studies relying on time-fixed effect models because both factors are time-varying.

3 We replicated Eq.1 controlling for year- and city-based price consumption index variations relative to city-based price consumption index variations and observed similar findings (results are available in Table S1 in Supplementary Materials).

